# Test-Retest Reliability of Hierarchical Proprioception Assessment of the Wrist

**DOI:** 10.64898/2026.08.07.26359977

**Authors:** Aravind Nehrujee, Kailynn Mannella, Robert W. Motl, Bruce Cohen, Milap S. Sandhu

## Abstract

Proprioception can be assessed in several ways, including movement detection, joint position matching, and matching across sensory frames of reference. These task types make different demands, yet they are rarely compared within the same participants on the same device, and psychometric data for wrist-focused batteries are limited. This work had two aims: to compare performance across different levels of proprioceptive judgment, and to establish the within-day test-retest reliability of each. We evaluated three robotic wrist tasks spanning judgments within a single reference frame and across reference frames: joint detection threshold (JDT), same-frame joint-to-joint matching (J-to-J), and cross-frame joint-to-visual matching (J-to-V).

**Methods:** Twenty neurotypical adults completed two identical sessions on the same day, separated by at least two hours, using a single-degree-of-freedom wrist robot. Outcomes were the kinematic detection threshold (degrees) for JDT and the mean absolute matching error (degrees) for J-to-J and J-to-V. Relative reliability was quantified with ICC (2,1) and 95% confidence intervals. Absolute reliability was quantified with the standard error of measurement (SEM) and the smallest detectable change at 95% confidence (SDC 95). Learning effects and differences across task levels were evaluated with paired t-tests or Wilcoxon signed-rank tests.

**Results:** ICC (2,1) was 0.959 [95% CI: 0.900 to 0.980] for JDT, 0.837 [0.640 to 0.930] for J-to-J, and 0.769 [0.500 to 0.900] for J-to-V. The %SEM ranged from 11.9% (J-to-J) to 15.6% (J-to-V). SDC95 was 0.85°, 1.71°, and 3.54° for JDT, J-to-J, and J-to-V, respectively. A small but significant practice effect was observed for JDT (S2 − S1: −0.16°, p = 0.033), but this was below the SDC95, and no learning effect was observed for J-to-J or J-to-V. We also observed that the absolute error increased monotonically across task levels, with all pairwise comparisons (JDT < J-to-J < J-to-V; all p < 0.01).

**Conclusions:** All three tasks demonstrated good-to-excellent within-day relative reliability. Error scaled with the computational demand of each task, with the largest errors observed for the cross-frame task, which required a transformation between the visual and joint reference frames. The reported SDC95 values provide task-specific thresholds for distinguishing measurement noise from true change in future intervention studies. Inter-day reliability and validation in clinical populations are the next steps.

## 1. INTRODUCTION

Proprioception, defined as the awareness of the mechanical and spatial state of the body and its musculoskeletal parts[1], is critical for planning, executing, and correcting motor actions[2]. When these signals are disrupted in several neurological conditions, as occurs in stroke[3], multiple sclerosis[4], and Parkinson’s disease[5], physical function declines. Therefore, accurate measurement of proprioceptive deficits is essential for understanding and treating them[6].

Proprioceptive signals, generated primarily by muscle spindles, with contributions from cutaneous and joint receptors and central motor commands, allow us to perceive the position and movement of our body[7]. Golgi tendon organs also signal the force generated by our muscles[8]. Importantly, proprioception is not determined solely by peripheral receptor input; rather, its perception depends on how afferent signals are interpreted within central body representations and integrated with visual, tactile, and vestibular information to estimate the body’s position and movement in space[9]. This suggests that proprioceptive performance may vary depending on the task’s frame of reference and the extent to which sensory information is transformed across reference frames[10] [11], [12].

Héroux et al.[13] proposed a framework for organizing proprioceptive assessments based on the frame of reference in which the judgment is made. **1**) **Low-level judgments** are made within a single frame of reference and include the detection, discrimination, and matching of proprioceptive stimuli. **2) High-level judgments** are made across different frames of reference and include localizing a body part in external space and matching across visual and joint frames, each of which requires a coordinate transformation. For example, reproducing a visually cued target angle with an unseen joint requires converting a visual representation of the target into a joint-space command [14]. Because high-level tasks involve additional processing, they are expected to impose greater computational demands and to produce larger errors than judgments made within a single frame.

Proprioceptive assessments in the clinics primarily sample the low-level portion of the framework [6], [11], [12]. A judgment made entirely within the joint frame, with vision occluded, isolates proprioceptive acuity. Still, it cannot reveal whether a person can transform information between the joint and visual frames. Bernard-Espina et al.[9] argued that many deficits attributed to proprioception after stroke instead reflect impaired cross-reference processing, the transformation of proprioceptive signals into other frames such as the visual frame, and that the two abilities can be selectively impaired. Because vision-based compensation depends on this transformation, pairing a within-frame judgment with a cross-frame judgment can begin to separate proprioceptive acuity from the ability to transform between frames, which a single within-frame test cannot do. A recent systematic review of 56 studies reported that 97% of proprioceptive assessments in clinical populations relied solely on low-level judgments, with virtually no assessment of high-level, cross-frame proprioception[15]. We therefore included both within-frame and cross-frame matching judgments.

Clinical proprioceptive assessment has traditionally relied on ordinal tools that rely on rater judgment and are too coarse to detect small differences or to distinguish sub-modalities[16]. Robotic devices, by contrast, deliver controlled stimuli and record high-resolution kinematics, enabling single- and cross-frame judgments to be quantified on the same device[17], [18], [19], [20], [21], [22]. The wrist is functionally central to hand use yet comparatively underexamined, and prior studies have focused primarily on one or two low-level sub-modalities[15].

For a battery of this kind to track proprioceptive change, whether over the course of recovery or in response to an intervention, an observed change must be distinguishable from measurement error[23], [24]. This requires task-specific estimates of reliability and the smallest change that exceeds measurement noise, neither of which is currently available for an assessment battery spanning these task types. Establishing these properties in neurotypical adults is the necessary first step before the battery can be applied in clinical populations. Proprioceptive judgments can be probed with either passively imposed or actively generated movements, and the two are not equivalent. Active movement yields more precise position estimates than passive movement for both joint angle and limb endpoint position[25], and active and passive judgments dissociate in absolute error, indicating that they do not draw on identical processes[26], [27]. This advantage is attributed to the efference copy of the motor command and to alpha-gamma coactivation[26], which are available only when the movement is self-generated. We therefore based the matching tasks on active reproduction.

Accordingly, the objective of this study was to evaluate the within-day test-retest reliability of three robotic proprioceptive tasks in neurotypical adults: a passive movement detection threshold (JDT), active same-frame joint-to-joint matching (J-to-J), and active cross-frame joint-to-visual matching (J-to-V). Together, these tasks span judgments made within a single reference frame and judgments made across reference frames[13]. We quantified relative reliability using the intraclass correlation coefficient (ICC) with 95% confidence intervals (CIs), and absolute reliability using the standard error of measurement (SEM) and the smallest detectable change at 95% confidence (SDC95). We also tested whether performance changed systematically between sessions and whether errors differed across the three tasks.

## 2. METHODS

### 2.1 Participants

Twenty neurotypical adults were recruited from the community and from research volunteer pools at Shirley Ryan AbilityLab and Northwestern University (Chicago, IL, USA). All procedures were completed within a single visit comprising two sessions separated by at least two hours to allow recovery from transient fatigue and attentional demands while maintaining a within-day test-retest design.

Inclusion criteria were age ≥ 18 years, ability to understand and follow task instructions, normal or corrected-to-normal vision, pain-free active wrist flexion-extension through a functional range, and intact cognition verified with the blind version of the Montreal Cognitive Assessment[28] (blind MoCA, score ≥ 18; maximum 22). Exclusion criteria were any history of neurological, orthopedic, or rheumatologic condition affecting the tested hand, wrist, or forearm; current upper-limb pain or injury; prior surgery likely to limit wrist range of motion; and current use of medications known to affect sensorimotor performance substantially.

All participants provided written informed consent before participation. The study was approved by the Northwestern University Institutional Review Board (STU00221436) and was prospectively registered at ClinicalTrials.gov (NCT06390930).

### 2.2 Apparatus

Assessments were performed using PLUTO (Plug-and-Train Robot) [29], a single-degree-of-freedom wrist robot that allows wrist manipulation in the flexion and extension axes. The robot is fully backdrivable for active movements and supports closed-loop position control for passive positioning. Joint angle was measured with a rotary incremental encoder (MILE 512-6400 CPT, 2 channels, Line Driver RS 422; Maxon Motor AG, Sachseln, Switzerland) at a resolution of 0.056 degrees. Task logic, stimulus presentation, and data logging were implemented in Unity 2022 (Unity Technologies, San Francisco, CA, USA). Participants responded with a handheld momentary button or a keyboard. Full hardware and control details of PLUTO have been described previously[29].

### 2.3 Experimental Setup

Participants sat in an armless chair with the trunk upright, the shoulder in no more than 15° of abduction, and the elbow flexed to 90° ± 10° (Figure 1). The tested forearm was secured to a padded support with proximal and distal adjustable straps to minimize translation and out-of-plane motion. The fingers were held in relaxed extension with the thumb in neutral. The robot’s rotational axis was aligned with the anatomical wrist flexion-extension axis using adjustable mounts, and alignment was verified by delivering small passive wrist oscillations and confirming the absence of forearm translation. Active range of motion (AROM) was measured at this position, and software limits for all tasks were set to ±60° around neutral. A rigid shroud occluded vision of the wrist and hand throughout all tasks. For tasks requiring visual feedback, the wrist angle was displayed on an eye-level monitor as a rotating line that mapped linearly to the wrist flexion angle at a 1:1 gain (Figure 1).

**Figure 1:**
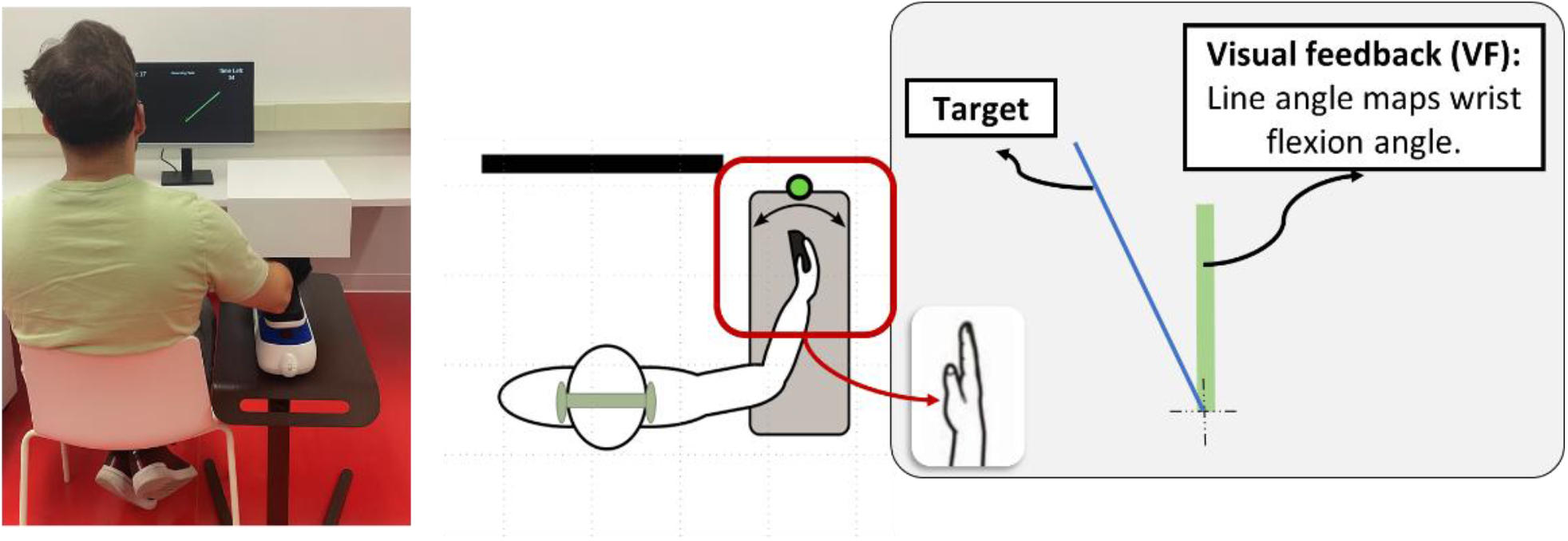
Experimental setup and visual feedback display. Left: participant seated at PLUTO with the forearm secured and vision occluded. Center: schematic showing forearm fixation and end-effector alignment to the wrist flexion-extension axis. Right: visual feedback display used during familiarization and position matching tasks; the rotating line maps linearly to wrist flexion angle, and the static green bar denotes the target.

### 2.4 Assessment Battery

The battery comprised two setup tasks (AROM measurement and familiarization with the visual display) followed by three proprioceptive tasks: The three proprioceptive tasks were administered in the same fixed order, J-to-J, J-to-V, and JDT, across all participants and both sessions to standardize task administration and minimize procedural variability.

JDT and J-to-J require judgments within a single reference frame, whereas J-to-V requires a judgment across the visual and joint reference frames. All targets and movements were confined to each participant’s AROM. Each task is detailed below. A video demonstration of the full battery is provided as Supplementary Video S1.

#### 1. Active range of motion (AROM)

Participants performed three voluntary wrist flexion-extension movements to their maximum comfortable limits of flexion and extension. The range of motion was averaged across the three repetitions. The start position for all subsequent tasks was set to the participant’s maximum active extension limit or 60°, whichever was lesser.

#### 2. Familiarization Task (point-to-point reaching)

Participants performed a point-to-point reaching task in which continuous wrist flexion-extension drove the cursor toward successive targets. A green rotating line (the cursor) mapped the wrist’s angular position to an on-screen rotation, such that wrist flexion and extension rotated the cursor accordingly; this provided visual feedback (VF) of wrist position (Figure 2). A blue line indicated the target angle, displayed at one of several rotations within the participant’s range of motion (Figure 2). From the start position, participants pressed a button to initiate each trial (Step 1). A target then appeared at an angular distance sampled from a uniform distribution between 25% and 35% of AROM-flexion relative to the current cursor position, with direction randomized while remaining within the software limits. Participants moved the cursor toward the target (Step 2) and held it within the target window for 300 ms to complete the trial (Step 3). Targets not reached within 3 s were replaced with a new target (Step 4). Participants completed three 60-second blocks. No outcome measures were derived from this task; this was done to help the participant understand the mapping between their wrist and the cursor.

**Figure 2.**
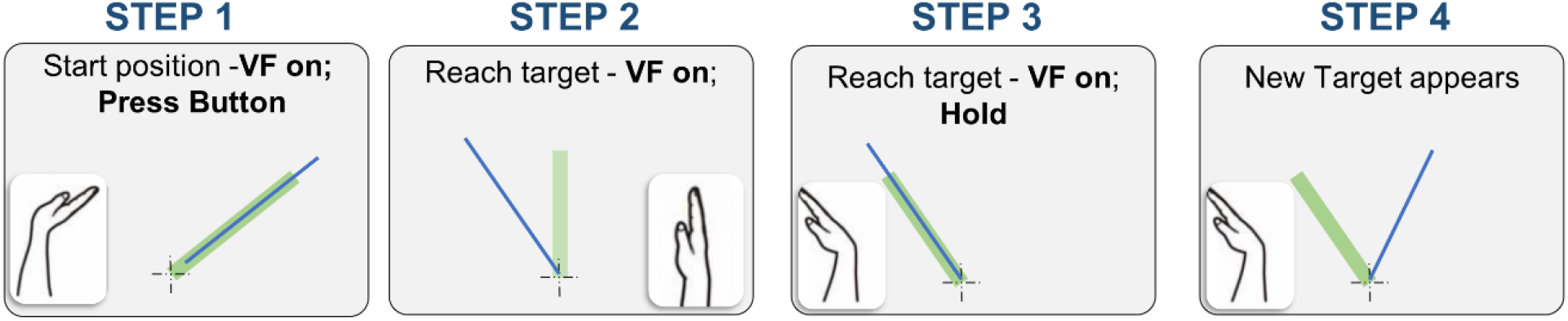
Familiarization task (point-to-point reaching). Participants moved a cursor, driven by wrist flexion-extension, to pseudo-randomly presented targets within their AROM. Step 1: start position with visual feedback on; button press initiates the trial. Step 2: The cursor moved toward the target with visual feedback on. Step 3: cursor held within the target window for 300 ms. Step 4: new target appears.

#### 3. Passive movement detection (joint detection threshold, JDT)

This task assessed the detection of a proprioceptive input, the simplest form of proprioceptive judgment within a single reference frame. Movement detection threshold is among the most widely used clinical measures of proprioception: the joint is moved passively at a constant velocity until the participant detects the onset of motion, providing a measure of peripheral proprioceptive sensitivity with minimal cognitive demand.

##### Procedure

Passive movement detection was assessed with a single-interval reaction-time paradigm. At the start of each trial, participants moved the wrist to the midpoint of their AROM, guided by on-screen text instructions. Once positioned, a fore period of 500 to 2,500 ms was introduced, drawn from a uniform distribution to minimize anticipatory responses. PLUTO then imposed a constant-velocity passive wrist rotation of 0.5°/s toward either flexion or extension, with direction randomized across trials. Participants were instructed to maintain complete muscular relaxation and to press a handheld button immediately upon perceiving the onset of motion. After the response, participants confirmed the perceived direction by pressing a left or right key. All rotations were delivered within the participant’s AROM. Trials were excluded if the participant responded before motion onset or reported an incorrect direction. Five flexion and five extension trials were collected in a randomized order per session.

##### Outcome measure

JDT (degrees) was computed on each trial as the angular displacement from motion onset to the response button press. The session score was the mean JDT across valid flexion trials. Extension trials were collected but excluded from scoring to maintain consistency with the matching tasks, in which all targets were defined within the flexion range.

#### 4. Joint position matching: same frame (joint-to-joint, J-to-J)

This task assesses low-level proprioceptive judgment made within a single reference frame. We chose ipsilateral matching within the same joint[25], [27], where participants actively moved the wrist to a target angle with visual feedback, memorized the position, and reproduced it without visual feedback using the same wrist. Both the encoding and reproduction phases occur within the joint reference frame[13].

##### Procedure

Each trial began with the wrist at the extension limit with cursor feedback; participants pressed a button to initiate the trial (Step 1). A target line appeared on the display; participants actively moved the wrist to align the cursor with the target (θ_target_) and pressed a button to confirm (Step 2, Figure 3). Cursor feedback was then removed, and participants returned the wrist to the start position; cursor feedback was briefly restored within ±5° of the start position to facilitate accurate repositioning (Step 3). In the reproduction phase, cursor feedback remained off; participants actively moved the wrist to the memorized angle (θ_repro_) within the same reference frame and confirmed their response by pressing a button (Step 4). Five target angles were used, set at 10, 30, 50, 70, and 90% of AROM-flexion, presented in random order with five repetitions per level (25 trials per session).

**Figure 3.**
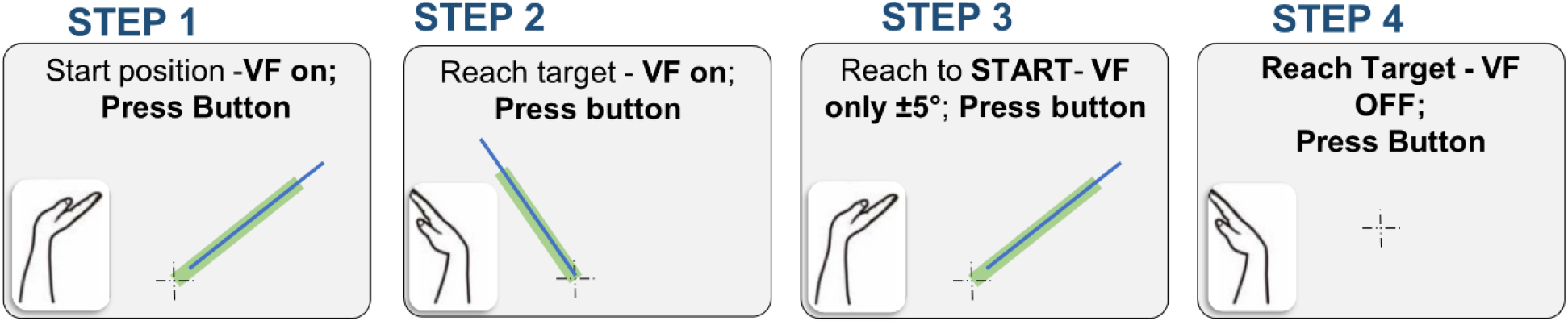
Joint position matching: same frame (J-to-J). Participants actively moved the wrist to a target angle with visual feedback, memorized the position, and reproduced it without visual feedback using the same wrist within the joint reference frame. Step 1: start in the start position with VF on; pressing the button initiates the trial. Step 2: participant moves the wrist to the target angle with VF on and confirms with a button press. Step 3: participant returns to the start position, with VF restored to within ±5°, and confirms with a button press. Step 4: participant reproduces the memorized angle without VF and confirms with a button press.

Returning the wrist to the fully extended start position (the maximum active extension limit) before each trial imposed a consistent movement history on the wrist muscles. Because muscle spindle output is history-dependent, this consistent conditioning standardizes the spindles’ thixotropic state and reduces trial-to-trial variability from this source[30], [31]. The same return to full extension preceded each trial in the J-to-V task.

##### Outcome measure

Absolute error (AE, degrees): *AE* = |θ_repro_ − θ_target_ |, where θ_target_is the target angle, and θ_repro_ is the wrist angle at the confirmation button press during the reproduction phase. The session score was the mean AE across all 25 trials. Twenty-five trials were included to sample five target positions distributed across the participant’s flexion range, with five repetitions at each position, providing repeated estimates across the range while maintaining a feasible assessment duration.

#### Joint position matching: cross frame (joint-to-visual, J-to-V)

This task assesses high-level proprioceptive judgment made across two reference frames. Participants viewed a target wrist angle presented in the visual frame and reproduced it in the joint frame, without visual feedback. To do so, the brain must transform a visual representation of the target angle into a corresponding joint position, a coordinate transformation from the visual to the joint reference frame.

##### Procedure

Each trial began with the wrist at the extension limit with cursor feedback on; participants pressed a button to initiate the trial (Step 1, Figure 4). A target line was displayed on the gauge (θ_target_). No cursor feedback was available during this phase. Participants actively moved the wrist to the angle they judged to correspond to the displayed target (θ_match_) and confirmed their response by pressing a button (Step 2). Participants then returned the wrist to the start position; cursor feedback was briefly restored within ±5° of the start position to facilitate accurate repositioning (Step 3). Five target angles were used, set at 10, 30, 50, 70, and 90% of AROM-flexion, presented in random order with five repetitions per level (25 trials per session).

**Figure 4:**
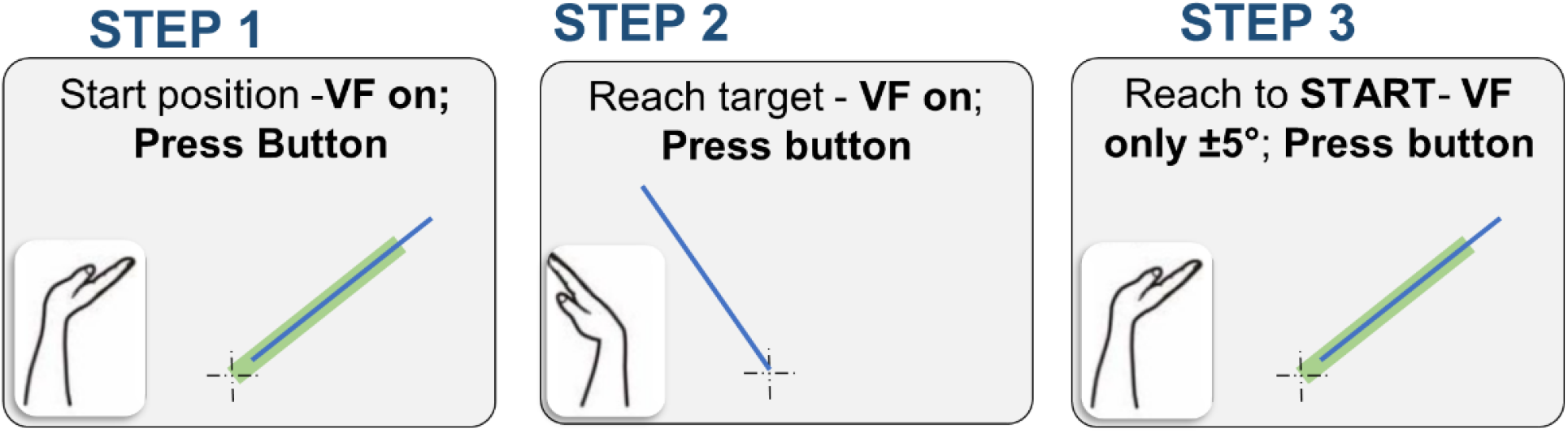
Participants viewed a target wrist angle on the display. They reproduced it without visual feedback using the same wrist, requiring a transformation from the visual to the joint reference frame. Step 1: start position with VF on; target line displayed; button press initiates the trial. Step 2: target disappears, and VF is removed; participant moves the wrist to match the displayed target angle and confirms with a button press. Step 3: participant returns to the start position with VF restored to within ±5°; a button press confirms repositioning.

##### Outcome measure

Absolute error (AE, degrees): AE = | θ_match_ − θ_target_| where θ_target_ is the nominal target angle, and θ_match_ is the wrist angle at the time of the confirmation button press. The session score was the mean AE across all 25 trials.

### 2.5 Protocol

Each visit comprised a fixed task sequence: active range-of-motion measurement, followed by the familiarization task, and then the three proprioceptive tasks, administered in the same order across both sessions (J-to-J, J-to-V, JDT). If participants did not demonstrate adequate task understanding during familiarization, the familiarization block was repeated before proceeding. Adequate task understanding was defined as completing at least 15 reaches within a 60-second familiarization block and confirming understanding of the wrist-to-cursor mapping. If either criterion was not met, the familiarization block was repeated before proceeding. The same examiner conducted both sessions.

### 2.6 Statistical Analysis

All analyses were performed in Python (NumPy, Pandas, SciPy, Statsmodels, Pingouin, Matplotlib). Session-level scores were computed from trial-level data as specified above and used for all inferential statistics. We assessed the normality of all session-level variables using the Shapiro-Wilk test.

#### Relative reliability

Relative reliability was estimated using ICC (2,1), a two-way random-effects model with absolute agreement[32], [33]. ICCs are reported with 95% confidence intervals and interpreted as poor (<0.50), fair (0.50-0.75), good (0.75-0.90), or excellent (≥0.9) [34].

#### Absolute reliability

Absolute reliability was quantified by calculating 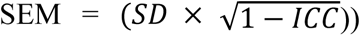 and SDC95 = (1.96 × √2 × SEM). Relative expressions (%SEM = 100 × SEM/Mean; %SDC95 = 100 × SDC95/Mean) were used as scale-free benchmarks[23]. The SDC95 provides a threshold that must be exceeded to conclude that an observed change reflects true modulation rather than measurement variability[35]. As benchmarks, %SEM < 10% was considered acceptable and 10-15% tolerable; %SDC95 ≤ 30% was considered acceptable and ≤ 10% excellent [36].

#### Learning effects

To evaluate practice effects, Session 1 and Session 2 scores were compared using paired t-tests when the within-participant differences were normally distributed and Wilcoxon signed-rank tests otherwise. Each task was analyzed independently, with no correction for multiple comparisons, because each assessment represented a separate hypothesis. Mean paired differences with 95% confidence intervals and Cohen’s d are reported.

#### Differences across proprioceptive task levels

To test whether proprioceptive error differed across task levels, we performed three pairwise comparisons using session-averaged scores. Because the within-participant differences were normally distributed for all comparisons, paired t-tests were used, with Holm-Bonferroni correction for multiple comparisons.

## 3. Results

Twenty neurotypical adults (19 right-handed) completed both sessions. Mean age was 30.8 ± 10.3 years (11Male/9 Female, 10 White, 6 Asian, 3 Black, 1 Hispanic). All participants reported full, pain-free wrist range of motion and no neurological, orthopedic, or rheumatologic history affecting the upper limb. A summary of all reliability indices is provided in Table 1.

**Table 1.** Test-retest reliability summary. JDT, joint detection threshold (degrees); J-to-J, joint-to-joint matching absolute error (degrees); J-to-V, joint-to-visual matching absolute error (degrees); ICC, intraclass correlation coefficient; SEM, standard error of measurement; SDC95, smallest detectable change at 95% confidence; %SDC95, SDC95 expressed as a percentage of the mean, p-value for session difference (Session 2 minus Session 1); each task tested independently.

|  | JDT | J-to-J | J-to-V |
| --- | --- | --- | --- |
| Mean, ° (SD) | 2.02 (1.55) | 5.20 (1.56) | 8.22 (2.57) |
| Range, ° | 0.25–7.57 | 3.25–9.45 | 2.91–13.66 |
| ICC (2,1) | 0.959 | 0.837 | 0.769 |
| ICC 95% CI | 0.90–0.98 | 0.64–0.93 | 0.50–0.90 |
| SEM, ° | 0.30 | 0.62 | 1.28 |
| %SEM | 15.1 | 11.9 | 15.6 |
| SDC95, ° | 0.84 | 1.71 | 3.54 |
| %SDC95 | 41.8 | 32.9 | 43.1 |
| Session 1, ° | 2.11 | 5.05 | 8.22 |
| Session 2, ° | 1.94 | 5.35 | 8.21 |
| S2–S1, ° | –0.16 | +0.31 | –0.01 |
| p | 0.033 | 0.131 | 0.983 |

### 3.1 Joint Detection Threshold

The mean JDT across both sessions was 2.02° (SD = 1.55°; 95% CI: 1.30° to 2.75°; range: 0.25° to 7.57°; Session 1: 2.11°, Session 2: 1.94°). One participant showed a notably elevated JDT relative to the group but demonstrated consistent performance across sessions (Figure 5). A small but statistically significant practice effect was observed for JDT (S2 minus S1: −0.163°, SD = 0.431°; 95%CI: −0.364 to +0.039°; Wilcoxon signed-rank, p = 0.033; dz = −0.38). However, the magnitude of this difference (0.16°) was well below the SDC95 of 0.85°, indicating that the shift was not clinically meaningful. The absolute and relative SEM were 0.305° and 15.1%, respectively. The SDC95 was 0.85° (41.8%). ICC analysis revealed excellent relative reliability (ICC (2,1) [95% CI] = 0.959 [0.900–0.980]).

**Figure 5.**
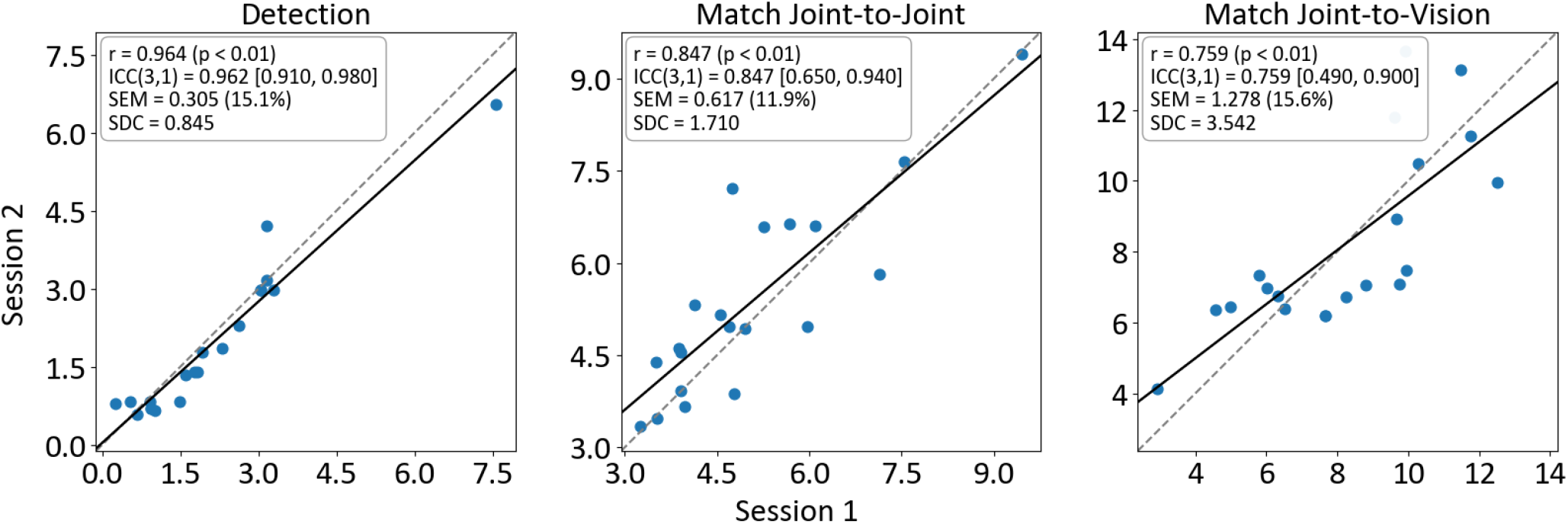
Test-retest reliability of wrist proprioceptive assessments. Session 1 versus Session 2 scores for joint detection threshold (JDT, left), same-frame joint position matching (J-to-J, center), and cross-frame joint position matching (J-to-V, right).

### 3.2 Joint Position Matching: Same Frame (J-to-J)

The mean J-to-J absolute error across both sessions was 5.20° (SD = 1.56°; 95% CI: 4.49° to 5.91°; range: 3.25° to 9.45°; Session 1: 5.05°, Session 2: 5.35°). We found no evidence of a learning effect between sessions (S2 minus S1: +0.308°, SD = 0.872°; 95% CI: −0.100 to +0.716°; paired t-test, p = 0.131; dz = 0.35) (Figure 5). The absolute and relative SEM were 0.617° and 11.9%, respectively. The SDC95 was 1.710° (32.9%). ICC analysis revealed good relative reliability (ICC (2,1) [95% CI] = 0.837 [0.640–0.930]).

### 3.3 Joint Position Matching: Cross Frame (J-to-V)

The mean J-to-V absolute error across both sessions was 8.22° (SD = 2.57°; 95% CI: 7.07° to 9.36°; range: 2.91° to 13.66°; Session 1: 8.22°, Session 2: 8.21°). We found no evidence of a learning effect between sessions (S2 minus S1: −0.009°, SD = 1.807°; 95% CI: −0.855 to +0.837°; paired t-test, p = 0.983; dz = −0.005). The absolute and relative SEM were 1.278° and 15.6%, respectively. The SDC95 was 3.542° (43.1%). ICC analysis revealed good relative reliability (ICC(2,1) [95% CI] = 0.769 [0.500–0.900]).

### 3.4 Differences across proprioceptive task levels

Proprioceptive acuity differed significantly across task levels, with error increasing monotonically from JDT (2.02°) to J-to-J (5.20°) to J-to-V (8.22°). Normality of all pairwise within-subject differences was confirmed by Shapiro-Wilk tests (all p ≥ 0.05), supporting the use of paired t-tests for all three comparisons. All three pairwise contrasts were significant after Holm-Bonferroni correction: JDT versus J-to-J (mean difference = 3.18°; paired t-test, p < 0.01; dz = 1.40), JDT versus J-to-V (mean difference = 6.19°; paired t-test, p < 0.01; dz = 2.27), and J-to-J versus J-to-V (mean difference = 3.02°; paired t-test, p < 0.01; dz = 1.50). Effect sizes were large across all comparisons, indicating that each task level reliably produced distinct error magnitudes in neurotypical adults.

**Figure 6:**
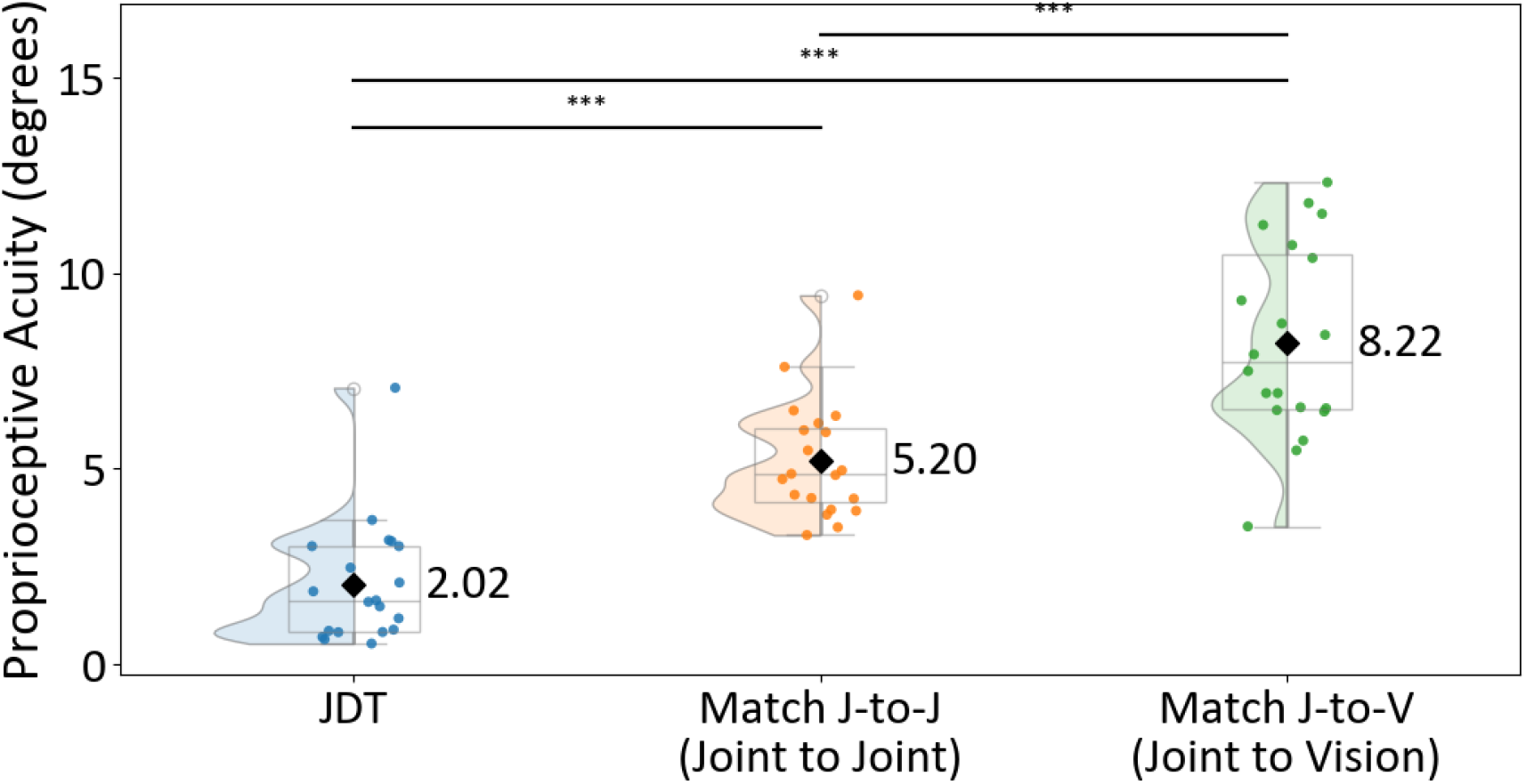
Proprioceptive acuity across task levels. Session-averaged scores for joint detection threshold (JDT), same-frame joint position matching (J-to-J), and cross-frame joint position matching (J-to-V) in all 20 participants. Diamond indicates group means. Horizontal bars indicate significant pairwise differences after Holm-Bonferroni correction (*** p < 0.01).

## 4. DISCUSSION

This study determined the test-retest reliability of three robotic wrist proprioceptive tasks in 20 neurotypical adults. The tasks were selected to span the low-to-high-level proprioceptive hierarchy proposed by Héroux et al. [2022]. All three tasks demonstrated good-to-excellent within-day relative reliability, with ICC values ranging from 0.769 for J-to-V to 0.959 for JDT. Absolute measurement error was task-specific, with SDC95 values of 0.85°, 1.71°, and 3.54° for JDT, J-to-J, and J-to-V, respectively, providing thresholds for distinguishing true change from noise in future studies. Performance was stable across sessions, with no session difference exceeding its task-specific SDC95. Proprioceptive error increased monotonically across task levels, with all pairwise differences significant, consistent with the greater computational demands imposed by cross-frame judgments.

### 4.1 Reliability of the measures

Within-day relative reliability was excellent for JDT (ICC = 0.959) and J-to-J (ICC = 0.837) and good for J-to-V (ICC = 0.769). ICC is a ratio of between-subject variance to total variance, and therefore depends heavily on the spread of scores in the sample tested. Because our participants were neurotypical adults with no proprioceptive impairment, they formed a relatively homogeneous group with narrow score distributions. This restricted between-subject spread compresses the ICC toward lower values, so the estimates reported here should be interpreted as conservative lower bounds. In a clinical sample in which proprioceptive deficits produce greater between-subject variability, ICC values would be expected to be higher.

Absolute reliability, which is not inflated by between-subject spread, is therefore the more directly interpretable index for a neurotypical sample. Measurement error (%SEM) ranged from 11.9% to 15.6% across tasks. The lowest measurement error was observed for J-to-J, in which participants reproduced an actively experienced joint position within the same reference frame. Higher error was observed for J-to-V and JDT, where a cross-frame coordinate transformation and a reaction-time response, respectively, introduced additional trial-to-trial variability. SDC95 estimates were 0.85° for JDT, 1.71° for J-to-J, and 3.54° for J-to-V. All three %SDC95 values exceeded the 30% acceptability benchmark [36]. This is partly a consequence of the restricted score range in a neurotypical sample: when mean scores are small, and participant variation is narrow, the absolute measurement error represents a larger fraction of the mean, inflating the percentage. The absolute SDC95 values, rather than their percentage expressions, are therefore the appropriate thresholds for use in future intervention studies.

Although JDT showed a statistically significant session difference (S2 − S1: −0.16°; Wilcoxon, p = 0.033; dz =−0.38), no session difference exceeded the SDC95 for its task. The JDT shift of 0.16° was well below the JDT SDC95 of 0.85°; J-to-J (+0.31°) and J-to-V (−0.01°) were similarly below their respective SDC95 values. Within-day performance was therefore stable across the battery, and a brief familiarization block was sufficient to support reliable measurement without further training. In clinical cohorts, greater between-subject variance from proprioceptive deficits would be expected to raise ICC values and lower %SDC95, bringing both within acceptable benchmarks. Confirming this requires empirical reliability estimates in clinical populations; until these are available, the absolute SDC95 values reported here provide the appropriate thresholds for future intervention studies.

### 4.2 Low- and High-Level Proprioceptive Judgments

Error increased monotonically across the three tasks, from JDT (2.02°) to J-to-J (5.20°) to J-to-V (8.22°), with large pairwise effect sizes. This ordering is consistent with the proposal that judgments requiring coordinate transformations across reference frames impose greater computational demands and produce larger errors than those made within a single frame[13]. The direction of the effect, specifically the larger errors observed for J-to-V than for J-to-J, is consistent with the long-standing observation that judgments across heterogeneous reference frames produce larger errors[37], [38], and with computational modeling evidence that motor biases during reaching arise primarily from systematic distortions in the transformation between visual and proprioceptive coordinate frames [39]. This pattern further supports the validity of inferences from scores as measures of proprioception.

It should be noted that the J-to-J task, while classified as same-frame, involves visual feedback during the encoding phase: participants align a visual cursor to a visual target before memorizing the position. The boundary between single-frame and cross-frame processing is therefore not absolute for this task. The proprioceptive memory carried into the reproduction phase was formed under visual guidance, potentially introducing a visual-to-joint transformation component during encoding. Future implementations could present the target as a passively imposed joint position to isolate proprioceptive encoding within a single reference frame more cleanly.

Inter-task correlations indicated that J-to-J and J-to-V were moderately correlated (r = 0.568, p = 0.009), whereas JDT was not correlated with either matching task (JDT versus J-to-J: r = −0.091, p = 0.702; JDT versus J-to-V: r = 0.124, p = 0.602) (Figure 7). This pattern suggests that detection and position matching tap distinct components of proprioceptive ability, and that same-frame and cross-frame matching share common variance while differing in the demand placed on coordinate transformation. Assessing multiple task types, therefore, provides complementary rather than redundant information about proprioceptive function.

**Figure 7.**
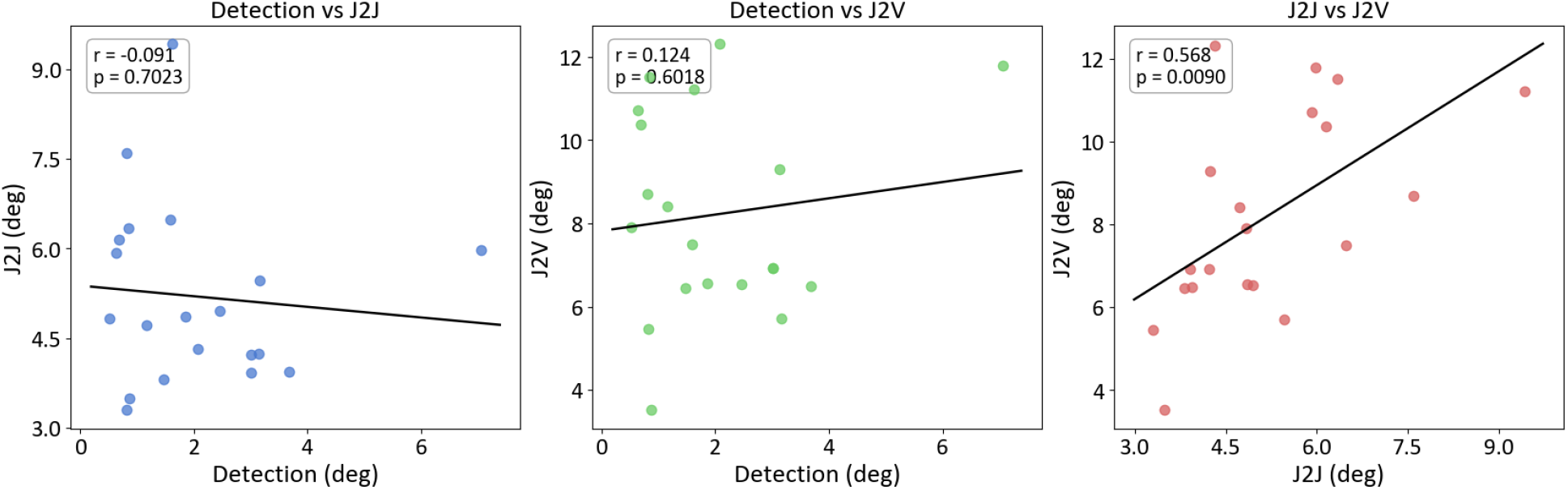
Inter-task correlations. Session-averaged scores for each pair of tasks in all 20 participants. Solid lines show the least-squares linear fit; Pearson r and p-values are inset. J-to-J and J-to-V were moderately correlated (r = 0.568, p = 0.009); JDT was not correlated with either matching task (r = −0.091 and 0.124; both p > 0.60).

These differences have potential clinical implications. Patients with neurological conditions such as stroke or multiple sclerosis may show disproportionate deficits on cross-frame relative to same-frame tasks, reflecting impaired coordinate transformation rather than peripheral proprioceptive loss alone. A recent systematic review of clinical proprioceptive assessment reported that 97% of studies relied on low-level judgments, with virtually no assessment of cross-frame proprioception[15]. A battery that samples both reference-frame structures, as assessed here, may therefore provide greater diagnostic specificity than low-level measures alone.

This ordering has been reported previously with different effectors and different stimuli. Héroux et al [14]. compared same-frame (grasp-to-grasp) and cross-frame (grasp-to-vision) judgments of grasped object width, and Robertson et al. [40] extended the comparison to the jaw; both studies found same-frame judgments to be more accurate and precise. Our results are consistent with these reports and extend them to the angle of a single joint, a judgment that does not depend on cutaneous signaling of object contact[14]. They also add a measurement dimension absent from that work. The task with the largest error here retained good reliability: J-to-V produced a mean absolute error of 8.22°, approximately 58% larger than J-to-J, yet its ICC (2,1) was 0.769, and its between-session difference was 0.009°. Under this account, a substantial portion of the error in a cross-frame task reflects a person-specific central calibration rather than trial-to-trial measurement noise. A stable but inaccurate calibration would produce large absolute error alongside high between-session agreement, which is the pattern we observed. A large J-to-V error should accordingly not be interpreted, on its own, as evidence of proprioceptive impairment.

### 4.3 Clinical Feasibility and Future Directions

Each participant completed the full battery in under 20 minutes, and the study established the reliability of the tasks in neurotypical adults; validation in people with proprioceptive impairments, such as stroke, multiple sclerosis, or peripheral neuropathy, is the logical next step. Future studies should establish inter-day and inter-week reliability, as within-day estimates may underestimate measurement variability across longer time intervals relevant to clinical monitoring. Studies in clinical populations should also examine whether the SDC95 values reported here are sufficiently small to detect the magnitude of proprioceptive change expected following intervention, and whether the J-to-V task provides additional diagnostic value over low-level measures alone.

A practical feature of this battery is that the participant’s own active movement drives both matching tasks. The robot here presents the targets and records wrist angle, but it does not move the limb during these tasks. The measurement, therefore, requires only accurate sensing of wrist motion, not actuation, which raises the possibility of administering the matching tasks without a robot. Wearable inertial sensors or markerless motion capture could, in principle, record the same active movements at lower cost and with greater portability [41], thereby broadening access beyond laboratories that own robotic devices. This does not extend to the detection task: JDT requires a controlled, passively imposed movement and therefore still depends on an actuator. Two conditions would need to be met before a sensor-based version could be adopted. First, the tasks require sufficient active wrist range of motion, which may limit their use in people with marked weakness, spasticity, or contracture. Second, wearable and markerless systems measure joint angles at a lower resolution than the robot’s encoder, so the SDC95 thresholds reported here would need to be re-established for the specific sensing platform before they could be applied.

### 4.4 Limitations

Reliability was established only in neurotypical adults; generalizability to clinical populations remains to be confirmed. Both sessions were conducted on the same day; inter-day and inter-week reliability have not been established. The battery assessed a single joint in a single plane, and generalizability to other joints is not implied. The sample size of 20 is consistent with recommendations for reliability studies [41], yields relatively wide ICC confidence intervals, particularly in the J-to-V task.

### 4.5 Conclusions

Three robotic wrist proprioceptive tasks spanning the low- to high-level hierarchy demonstrated good-to-excellent test-retest reliability in neurotypical adults. The SDC95 values provide task-specific benchmarks for distinguishing true change from measurement noise. Validation in clinical populations with proprioceptive impairments is a necessary next step.

## Data Availability

All data produced in the present study are available upon reasonable request to the authors

## ACKNOWLEDGEMENTS

The authors thank all participants for their time and commitment. We thank the members of the Synapse Lab at Shirley Ryan AbilityLab for their support throughout data collection, and the R2D2 group at IIT Madras and the BioRehab Group at CMC Vellore for making the PLUTO robotic platform available for this study. The authors gratefully acknowledge support from the National Multiple Sclerosis Society (NMSS; Grant RG-2307-42156).

## AUTHOR CONTRIBUTIONS

A.N. contributed to the conceptualization of the study, protocol design, data collection, and manuscript preparation. K.M. contributed to the data collection and manuscript revisions. M.S., R.M., and B.C. contributed to the study design, interpretation of findings, and revision of the manuscript. M.S. supervised and funded the project, contributed to the study design, interpretation of findings, and revision of the manuscript. All authors approved the final version and agree to be accountable for the work.

